# Deaths among COVID Cases in the United States: Racial and Ethnic Disparities Persist

**DOI:** 10.1101/2020.11.15.20232066

**Authors:** Madeleine Short Fabic, Yoonjoung Choi, David Bishai

## Abstract

Using COVID-19 Case Surveillance Public Use Data by the Centers for Disease Control and Prevention, we estimate monthly age-adjusted case fatality rates (CFR) for four major groups: non-Hispanic (NH) whites, NH Blacks, NH Asians, and Hispanics. Available data show that CFRs across race/ethnic groups have become more equal over time. Nevertheless, racial and ethnic disparities persist. NH whites consistently experience lower CFRs; NH Blacks generally experience higher case fatality among younger patients; and NH Asians generally experience higher case fatality among older patients. Age-adjusted CFRs reveal dramatically different racial and ethnic disparities that are hidden by crude CFRs. Such adjustment is imperative for understanding COVID-19’s toll.

## Introduction

Of all race/ethnic groups in the United States, Blacks, Hispanics, and Indigenous Americans experience the highest COVID-19 incidence and mortality.^1^ Less is known, however, about differential survival post-infection.^2,3^ Although age is the single most important factor for survival, case fatality rates (CFR) disaggregated by both age-and-race are rarely published. We report the extent to which racial/ethnic disparities in CFR remain after adjusting for age patterns among those infected, and how such disparities have changed over time.

## Methods

Using COVID-19 Case Surveillance Public Use Data by the Centers for Disease Control and Prevention (CDC), we estimate monthly age-adjusted CFR by race and ethnicity for four major groups: non-Hispanic (NH) whites, NH Blacks, NH Asians, and Hispanics.^4^ The November 3 release includes all COVID-19 cases reported to CDC through October 16—totaling 5,760,066 de-identified cases—alongside data on age (in 10-year groups), race/ethnicity, and survival status. We measure monthly CFR as the percent of cases reported in each month that have resulted in death either during the month or later. Eligible cases are those reported between 1 March and 31 August. We limit trend analyses to March through August, considering delayed reporting and clinical course of COVID-19. Reporting of race/ethnicity has remained poor (missing in 40.7% of cases), and we restrict our analyses to cases where both age and race/ethnicity were reported. We employ a direct standardization method to compare age-adjusted CFR by race/ethnicity, using age-specific CFR by race/ethnicity and standard age-distribution of cases, pooled from all cases in the data.^5^

## Results

CFRs—crude and age-adjusted—have declined over time across each racial and ethnic group (Figure 1). Age-adjusted CFRs reveal dramatically different racial and ethnic disparities that are hidden by crude CFRs. Adjusting for age-composition among cases reveals that NH whites have experienced the lowest CFRs, while NH Blacks, NH Asians, and Hispanics have experienced elevated case fatality. This finding reflects the pattern observed in age-specific CFRs: NH Blacks have generally experienced higher case fatality among younger patients and NH Asians have generally experienced higher case fatality among older patients (Figure 2). For NH Asians, elevated case fatality has been masked by COVID-19 mortality rates. Importantly, the magnitude of the CFR disparities has diminished over time.

**Figure 1.**
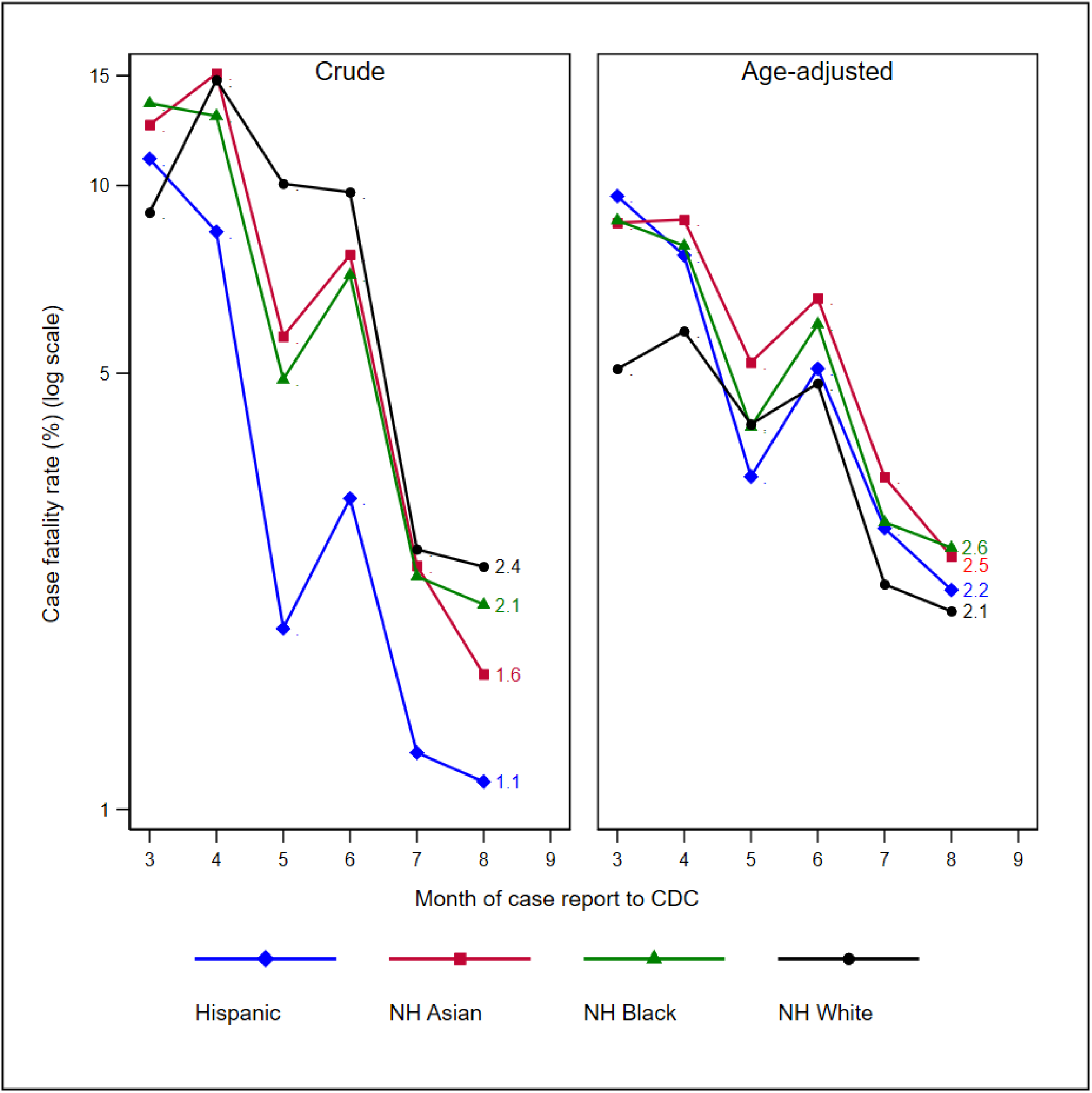
Trends of monthly case fatality rates among COVID-19 cases by race/ethnicity: crude and age-adjusted. NH: Non-Hispanic. Results for four largest single race/ethnicity groups are presented. Monthly case fatality rates refer to percent of cases reported in each month that have resulted in death either during the month or later.

**Figure 2.**
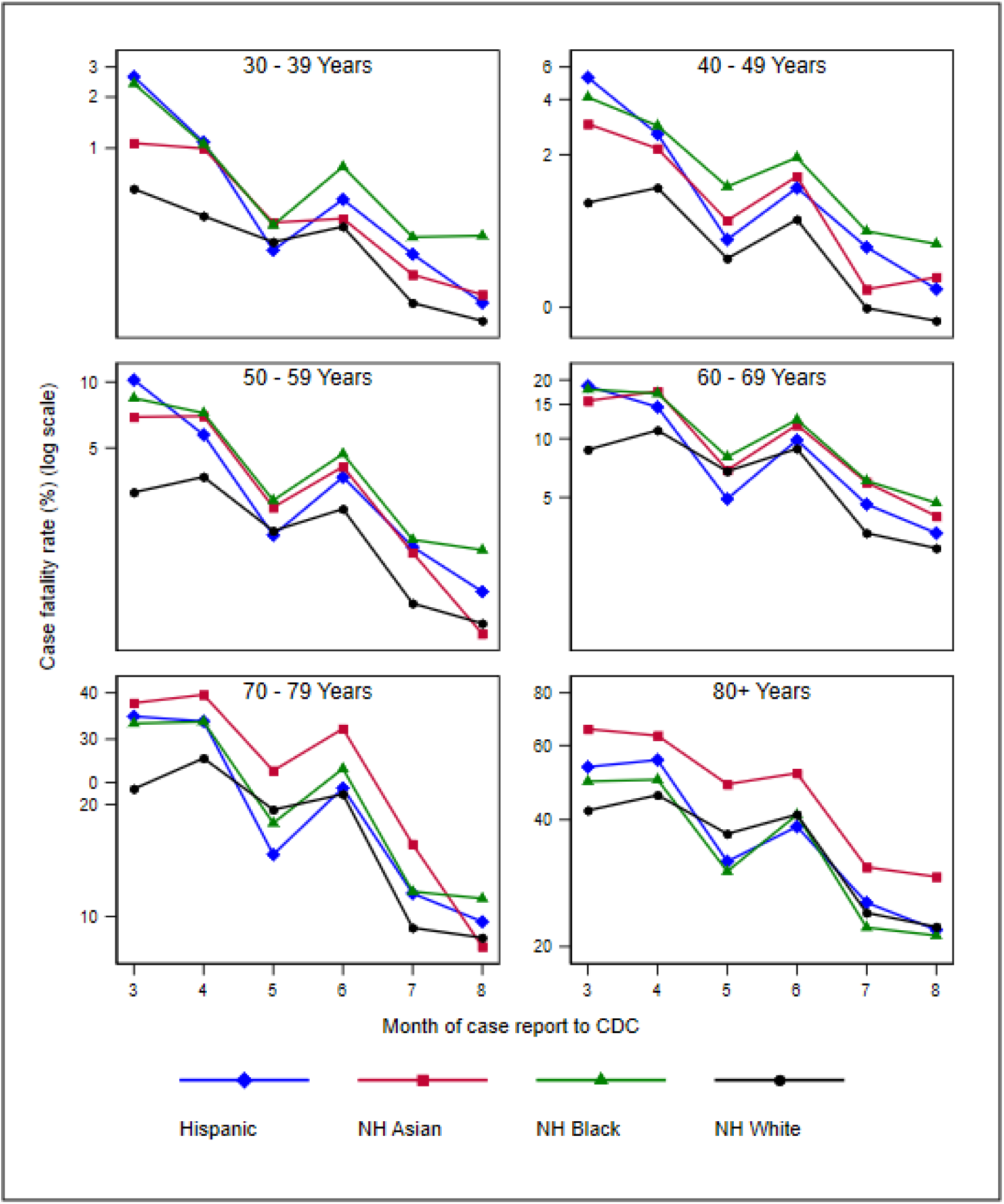
Monthly age-specific case fatality rates across race/ethnicity groups. NH: Non-Hispanic. Results for four largest single race/ethnicity groups are presented. Monthly case fatality rates refer to percent of cases reported in each month that have resulted in death either during the month or later.

## Discussion

This study is subject to at least three limitations. First, COVID-19 disproportionately impacts minorities, but without complete reporting of race and ethnicity, the full scope of inequities remains unknown. Second, the dataset does not include geographic variables (e.g. state). Without such information, we cannot ascertain whether the reduction in disparities reflects a geographical shift over time in where COVID is occurring or true changes within places that made CFR become more equal across race and ethnicity of a spatially defined population. Third, despite restricting our trend analyses to those cases reported in March through August, there is still a potential for a limited amount of right censoring in the ascertainment of death, which may artificially deflate CFRs for August.

As the pandemic has progressed, knowledge of COVID-19 has advanced, health providers’ experience managing COVID-19 has grown, and CFRs have fallen. Subject to its limitations, available data show that CFRs across race/ethnic groups have become more equal. The narrowing of case fatality disparities could be the result of more equal extension of bio-medical therapeutic advances across racial and ethnic groups, which would signify that health care systems in the US have improved their ability to equitably respond. Nevertheless, racial/ethnic disparities persist. To eliminate such COVID-19 inequities, public health, healthcare, and research communities must immediately and continuously recognize and dismantle the systemic and institutional racism that perpetuates racial and ethnic health disparities.^6^

## Data Availability

Data are available online via CDC’s “COVID-19 Case Surveillance Public Use Data.”
The study is reproducible; data management and analysis code in R Studio is available to the public.

https://data.cdc.gov/Case-Surveillance/COVID-19-Case-Surveillance-Public-Use-Data/vbim-akqf

